# Temporally Corrected Dose Accumulation – Next Steps in the Biology of Reirradiation

**DOI:** 10.1101/2024.08.25.24312201

**Authors:** Vishhvaan Gopalakrishnan, Bezhou Feng, Eashwar Somasundaram, Julia Pelesko, Kevin Stephans, Anthony Magnelli, Shlomo Koyfman, Gregory Videtic, Peng Qi, Xiaofeng Yang, Elliot M. Abbott, Jonathan W. Piper, Richard L.J. Qiu, Jacob G. Scott

## Abstract

**Introduction:** Treatments encompassing multiple courses of radiation are becoming increasingly common in the management of oligometastatic disease, offering opportunities to extend progression-free and overall survival. However, a major challenge in clinical practice is the lack of standardized methods to assess and mitigate toxicity risks associated with successive radiation treatments. Furthermore, normal tissue recovery post-radiation remains poorly characterized, and the absence of standardized documentation for radiotherapy history complicates large-scale research efforts. To address these limitations, we propose the development of a novel DICOM-compatible object for integration into patient medical records.

**Materials and Methods:** We generated software designs and bundle mathematics that demonstrate the utility of this DICOM object and how various dose forgiveness algorithms can be applied to the data. We include simple linear, exponential, logarithmic, and Gaussian recovery algorithms as well as complex non-linear algorithms based on the literature currently available.

**Results:** We applied the tool to an anonymized patient dataset, demonstrating the mathematical analysis applied to the data found in the new DICOM object. Noting ease and efficacy, we demonstrated that, in contrast to the current practice of gathering and structuring information distributed across electronic medical records, ready access to prior radiation courses accomplished two goals. (1) Facilitate data collection and analysis by streamlining access to comprehensive radiotherapy history, enabling researchers to conduct large-scale studies, and ultimately improve our understanding of tissue recovery. (2) Enhance clinical decision-making by enabling clinicians or software tools to leverage this data to personalize treatment plans, support clinical decision making to minimize toxicity risks during re-irradiation. For the anonymized patient, our analysis demonstrates safer delivery of re-radiation plans when viewed in the lens of dose forgiveness.

**Conclusions:** A novel DICOM object which keeps track of radiation treatments enables clinicians to factor tissue recovery and response into planning safer multiple radiation therapy courses and facilitates cross-institution research on re-irradiation and dose forgiveness.

## 1 Introduction

Re-irradiation is becoming increasingly prevalent in clinical settings for treatment of recurrent and oligometastatic or oligoprogressive disease. With evolving treatment paradigms for cancer leading to better prognoses for those with widespread disease, oligometastatic (i.e. limited in location and number) and widely disseminated metastatic disease are increasingly on a dynamic spectrum^1^. In addition to novel systemic agents, clinical trials have shown evidence that stereotactic body radiation therapy (SBRT) can extend progression-free and overall survival in patients with oligometastatic disease^2,3^. When planning radiation therapy (RT) courses that may involve treating individual sites in an organ that develop over time (metachronous lesions), planning new RT courses is affected by courses of previous radiation. Importantly, this requires striking a balance between the therapeutic effect of a local therapy, such as SBRT, and its potential injury to the organs-at-risk (OARs). For SBRT in particular, dose constraint tables developed by Timmerman, and utilized in cooperative group trials and recommendations such as AAPM Task Group 101, are widely considered to be the “gold standard” for describing radiation dose limits for different OARs above which the risk of developing tissue complications significantly escalates^4^.

Another well-studied approach to estimating treatment safety for single courses of radiation therapy is the Lyman-Kutcher-Burman (LKB) model, which calculates toxicity probability as a function of radiation dose received by an organ. This model has successfully predicted the toxicity from conventional fractionated RT to multiple OARs but has also shown to be unsuccessful in predicting toxicity in some cases, e.g. SBRT^5^. The accuracy of the LKB model relies heavily on the selection of parameters, and the use of a wide range of parameters means that the results must be interpreted with caution^6^. Alternative models exist to predict tissue toxicity; however, these models were developed with relatively limited data, so their accuracy also remains to be validated^7^.

For sequential SBRT regimens, one current commonly adapted strategy to assess the safety of administering multiple RT courses involves the calculation of a cumulative dose map. To generate this map, prior radiation doses are superimposed onto the upcoming SBRT plan using image registration techniques. When accumulated dose calculations suggest OARs could potentially receive a total dose exceeding commonly accepted constraints, clinicians must decide whether to alter the RT dose and potentially compromise local tumor control or to proceed treating with an increased risk of toxicity. When considering such choices, some radiation oncologists accept that with sufficient time between radiation courses, some radiation dose can be “discounted” or “forgiven”^8^. This comes from prior animal studies demonstrating some normal tissue recovery and a higher tolerance for radiation delivered over two courses separated by time^7,9^.

With this in mind, we foresee clinical scenarios where treatment with multiple courses of radiation will become increasingly common, especially in light of recent work directed at better understanding the effect of oligometastatic-directed therapy on patient survival^10,11^. As such, physicians need a way to consistently and reliably account for dose accumulations over multiple courses of radiation, in order to and to reproducibly estimate normal tissue recovery. To achieve this, we propose developing a reference Digital Imaging and Communications in Medicine (DICOM) object that represents a virtual model of the patient, onto which all radiation plans are mapped at the voxel level at the time of treatment completion. This tool, together with mathematical models of RT dose forgiveness over time, will allow clinicians to factor tissue recovery and response into the planning of multiple RT courses, as well as to collect data on the efficacy and toxicity of specific treatment plans. The models of dose “forgiveness” can be as conservative as simple arithmetic addition of doses across treatments, or as complex as combining patient specific genomic data^12^ with tissue-specific recommendations from the literature. Given the inherent uncertainty of such modeling, the tool is designed to provide structured guidance to help radiation oncologists standardized their approach to understanding the risks and benefits of re-irradiation.

Despite the dearth of data, some guidelines do exist for repeat radiation treatment. One review posits that for spinal cord SRS, re-irradiation EQD2 should not exceed 25 Gy and cumulative EQD2 should not exceed 70 Gy^9^. However, the authors also note the limited case control data available and write that these guidelines are likely conservative. A meta-analysis of patients with recurrent rectal cancer found that re-irradiation using total cumulative doses ranging from 66.4 to 103.3 Gy was well tolerated, with acute and late grade 3-4 toxicity rates of 11.7% and 25.5% respectively^13^. SBRT re-irradiation for rectal cancer appears especially promising in eligible candidates, with one study showing high response and low toxicity using a 30 Gy dose in 5 fractions^14^. A broader review examined re-irradiation across multiple tissue types and concluded that re-irradiation is feasible using either a hyperfractionated or stereotactic approach^15^. Given the high threshold for cumulative dose without toxicity in a majority of patients, these data indicate that dose forgiveness is a function of both the underlying tissue and cancer type (and likely individual patient biology).

## 2 Models and Methods

The first step of creating temporally corrected dose accumulation (TCDA) plans begins with selecting the model of tissue recovery to use. If providers assume no dose recovery and use arithmetic addition of doses across treatments, subsequent radiation doses are quickly hampered the maximal tolerable limits. Given the proven survival benefit of treating oligometastatic lesions with SBRT^2,3,16–18^, this conservative assumption will often limit our ability to continue treating if multiple courses are required, even if the patient may tolerate the treatment. In one study, primates were able to tolerate an extra 37.2 Gy EQD2 (equivalent dose in 2 Gy fractions) of cumulative radiation before toxicity than would be predicted by no recovery over an intervening two years^19^. Clinical data also indicate surprising resilience of spinal cord tissue to radiation myelopathy upon re-irradiation of tissue.^20,21^ Even with these studies, a precise way to model the RT dose forgiveness in the spinal cord, let alone other tissues, is not clear. This uncertainty makes rigorous study of radiation toxicities difficult. Our tool was created with the aim to expose these models of dose forgiveness to the end user, allowing them to compare and contrast between level of forgiveness and finely explore tissue toxicities with each model.

To this end, simple mathematical decay functions including linear, exponential, logarithmic, and Gaussian decay are shown in the software for consideration. In reality, a more complex non-linear decay function that combines tissue type, genomics, age, and other patient characteristics can be investigated to fully characterize dose forgiveness. By combining dose decay models with dose limits such as those proposed from Quantitative Analysis of Normal Tissue Effects in the Clinic (QUANTEC), we create a robust model that computes the maximum tolerable dose of a planned RT course as a function of tissue, time, and dosage of prior radiation therapies. However, gathering sufficient data to develop such a model is not feasible without a standardized way to compare and collate plans for patients treated in different institutions and over time. The genesis of this new data standard that stores a patient’s temporal radiation history and a 3D “body map” aims to solve quintessential dilemma in clinical radiation oncology.

Both the treatment planning image and radiation history can be represented using the DICOM medical imaging standard through a pre-existing entity within the standard^22^. Over time, the standard has seen numerous iterations and has accommodated radiotherapy information since 1997, with the publication of Supplement 11 by Working Group 07. In 2016, Working Group 06 created a second generation of RT objects in Supplement 147, including “RT course” which is a top-level entity that encapsulates both an RT prescription and physically delivered doses for one or more tumors^23^. We propose storing each round of RT that a patient receives as a separate RT course. After each fraction of radiation, we update the current course with the delivered dose and apply image registration to map the dose into corresponding voxels (a three dimensional volume) at specific locations in the underlying virtual model of the patient. The parameters of the image registration process will be stored separately as DICOM files and contain references to the treatment planning image and RT doses which were used in the mapping process. This separation provides flexibility in case the reference virtual model or preferred registration algorithm changes in the future. In a hypothetical situation, if a patient requires re-irradiation, the clinician is able to visualize temporally corrected dose accumulations of prior treatments by selecting from one of multiple algorithms which process the stored RT courses. Once these calculations are performed on the reference virtual model, the corrected doses are then mapped back onto the current treatment planning image and imported into a desired RT planning application. Therefore, we are able to leverage the sophisticated optimization algorithms that already exist. **Figure 1** gives examples of several decay functions that could be used initially.

**Figure 1.**
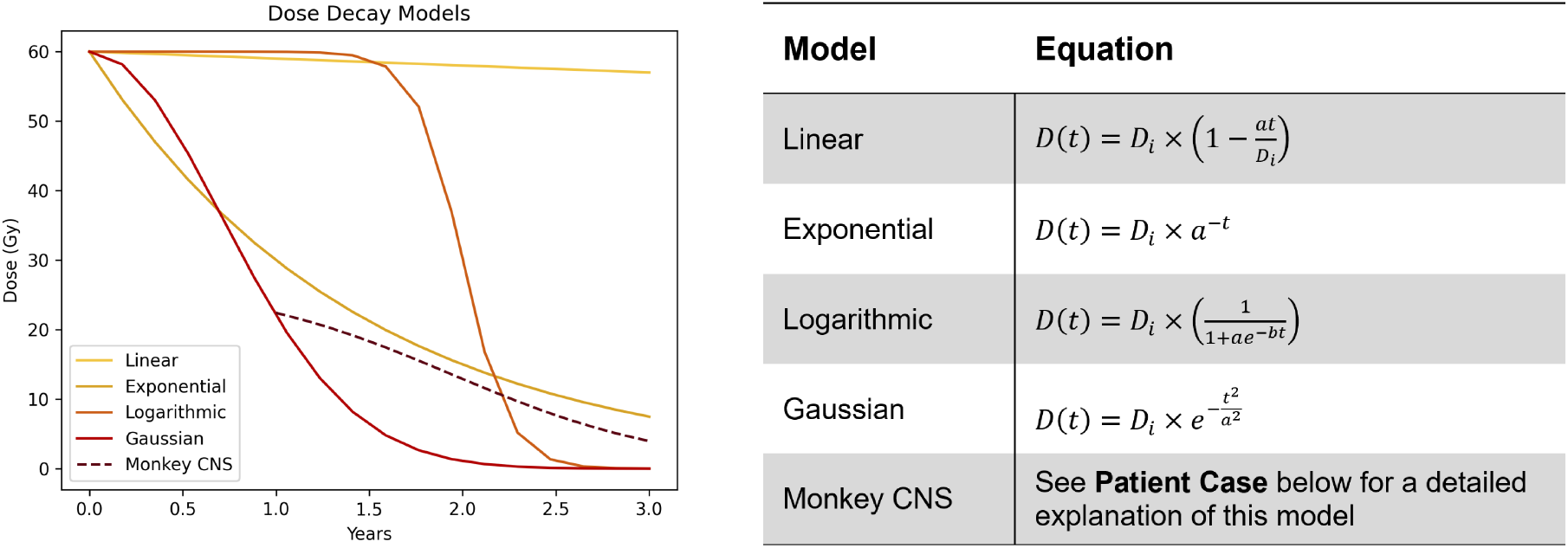
Examples of temporally adjusted radiation dose using different models of recovery. In the table of corresponding equations, *D*_*i*_ represents the initial dose, *t* represents time, and *a* and *b* are constants (chosen arbitrarily in the left plot).

The choice of re-irradiation will still be at the discretion of clinician, but such a feature would allow for a more quantitatively informed judgment of tissue recovery, and more rigorous study and comparison of plans and outcomes, and subsequent model refinement. Furthermore, our proposed standard is inherently extensible as the underlying RT courses will be stored for analysis by future algorithms. **Figure 2** shows what a future user interface tool might look like. We envision this tool to not only stand alone, as demonstrated, but also incorporated into existing RT planning or contouring software.

**Figure 2.**
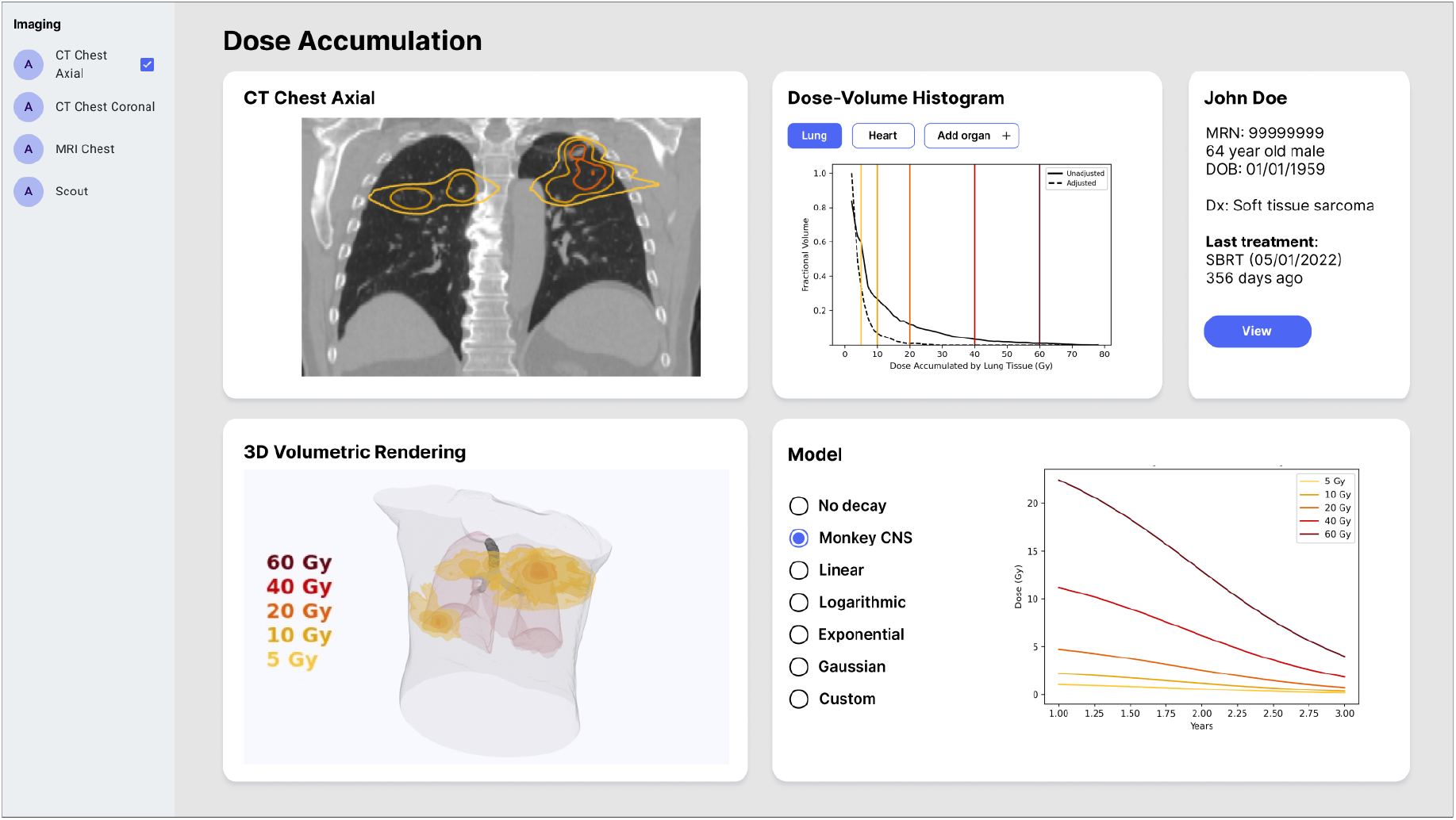
Proof of concept for a user interface that presents temporally corrected dose accumulation. The clinician has multiple options for visualization allowing them to tailor their radiotherapy planning.

Eventually, we envision that this new object in the medical record will store the algorithm used, radiation dosage given, tissue type irradiated, and any toxicity experienced.

## 3 Patient Case

Consider a sample patient, Mr. X, a man with oligometastatic soft tissue carcinoma. Mr. X had no significant past medical history and was in his usual state of health until being diagnosed with high grade myoepithelial carcinoma in the left distal thigh. His treatment entailed a large resection with neoadjuvant and adjuvant radiation therapy using a combination of external beam and brachytherapy, which successfully controlled the primary lesion. Nine months later however, he presented with multiple metachronous lesions in both lungs, which developed despite treatment with adjuvant systemic chemotherapy. These lesions were not apparent in his initial course and were treated with 10 sessions of SBRT over approximately two years. The SBRT was successful in controlling the targeted nodules; however, estimating the extent of tissue injury between treatments was challenging. In this scenario, a reference object that maps all of Mr. X’s therapies geospatially onto a reference 3D model might have allowed for easier appraisal of the safety of his treatment plan.

Even though the lung is a parallel organ (composed of many many subunits that mitage the effects of radiation to one part of the organ), serial SBRT treatment can often lead to lung doses that exceed tolerances. Furthermore, if a patient was treated by conventional radiation within their thorax, interplay between SBRT treatments and prior radiation will expose patients to increased risk of side effects to important organs at risk (such as the aorta, esophagus, and airway). Establishing dose forgiveness for these organs at risk are critical to establish if safe treatment can be administered to progressive disease.

**Figure 3** shows a proof of concept model using the summation of the first four of Mr. X’s SBRT treatment sessions. We use a subset of the full 10 sessions to simplify computation of the dose adjustment and for ease of visualization in the unadjusted condition. **Figure 4** further illustrates the impact of temporal correction by showing dose accumulation after each treatment. The selected SBRT sessions took place over the course of two years. Under the assumption that Mr. X’s lung tissue and spinal cord experienced recovery, let us now consider how we might plan for dose escalation in future SBRT sessions. For illustrative purposes only, if we use the previously described primate SRS dose toxicity data from Jones et al.^7^, we apply a model of temporal dose adjustment. The primate CNS model was chosen from the aforementioned theoretical models as a balance between optimistic and conservative tissue recovery rates.

**Figure 3.**
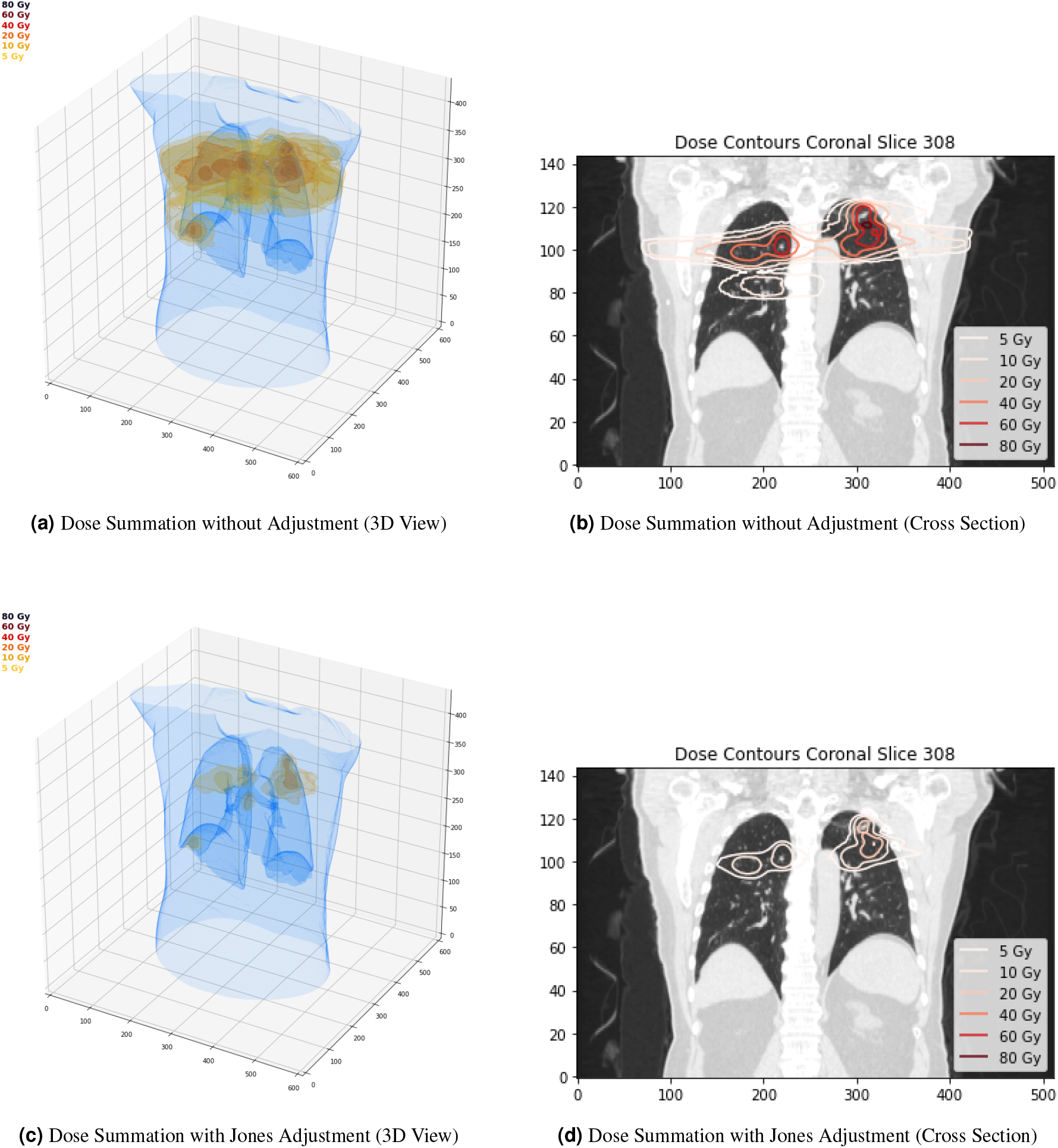
Visual proof of concept for dose adjustment. (a) The three dimensional view of summing four dose regimens of Mr. X’s SBRT treatment course without consideration of tissue recovery. (b) Demonstrates that with this approach, some of his lung tissue would have been exposed to toxic doses of radiation in the 80 Gy range. Using a dose adjustment equation described by Jones et al., we temporally correct the dose values based on time passage since treatment. With this equation, there is substantial reduction of accumulated dose shown in (c) and (d).

**Figure 4.**
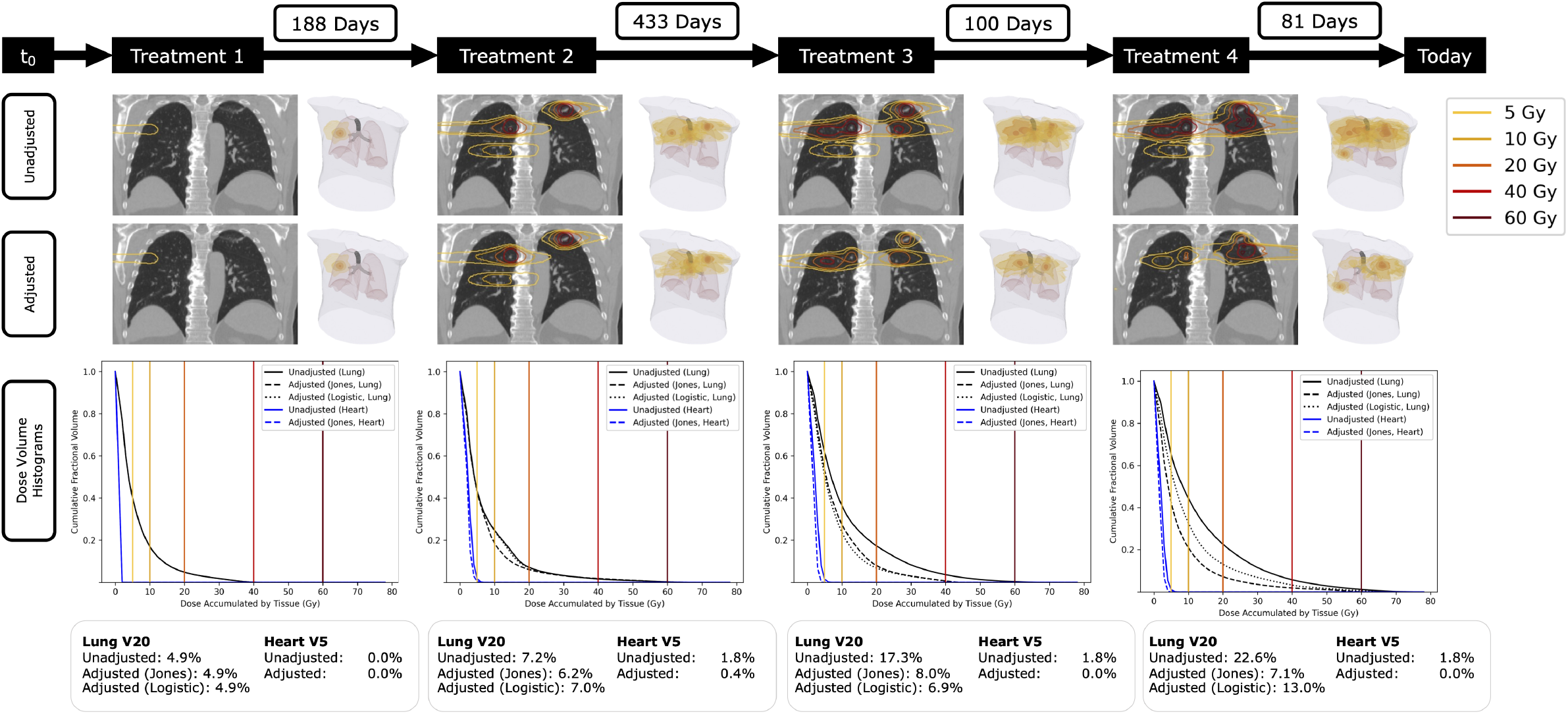
Timeline of dose accumulation in lung tissue with and without temporal adjustment. The first row shows cumulative radiation isodose lines after each treatment using arithmetic summation without adjustment. The second row shows the same tissue with doses corrected using the Jones et al. equation. The third row contains cumulative dose-volume histograms that compare the volume of lung and heart tissue exposed to a range of radiation doses with and without adjustment. Two different adjustments are used in the third row: the Jones et al. equation and a logistic decay function. The fourth row shows lung V20, the percentage of lung volume receiving *≥* 20 Gy, which is a commonly used metric for evaluating the risk for radiation pneumonitis. The fourth row also shows heart V5, the percentage of heart volume receiving *≥* 5 Gy, which is a commonly used metric for evaluating the risk of cardiac events due to radiation. Note that after treatment 4, both lung V20 and heart V5 are significantly higher in the unadjusted condition.

In this model, we first calculate what percentage of the maximum tolerable dose can be given after a recovery period using the equation:

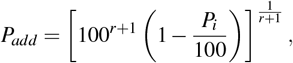

where *P*_*i*_ is the initial dose, *D*, as a percentage of the maximum tolerable dose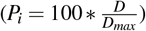. *r* is an experimentally derived constant that varies based on the time *t* since the initial dose:

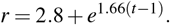

Because 100 *™ P*_*add*_ will equal the temporally adjusted *P*_*i*_, we can use the following equation to determine the numerical dose:

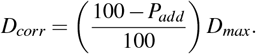

Thus, *D*_*corr*_ is the temporally corrected adjusted dose at each voxel which we use when evaluating our plan against established dose limits. We typically use 50 Gy for spinal cord tolerance and we can use this formula to determine the appropriate dose of radiation to administer to stay within this limit (even in the reirradiation setting).

**Figures 3 and 4** demonstrate how having a quantitative method to adjust and evaluate dose accumulation through time could help guide clinical judgement. With the increasing arsenal of chemo-, immuno-, and targeted^24^ therapies^25^, advances in radiation therapy, and development of new surgical techniques, physicians will be better equipped to provide individuals with personalized therapy regimens. With advanced therapies applied to patients, radiation oncologists are increasingly treating the same patient with multiple courses of radiation. In this setting, accurate communication between care providers is critical, especially if they extend across different institutions. The proposed storage of radiation treatment associated data in the DICOM object will allow for such synchronization. This approach leverages the existing standards published by DICOM to record chemotherapy history while allowing for future capabilities, such as indicating sites of surgical resections. We envision that with the addition of clinical data, including side effects, to the DICOM clinicians will better be able to understand a patient’s oncologic treatment history from a quick visual appraisal. This will be a much needed improvement of the current approach of searches through archives of manually written patient notes from multiple sources, by consolidating pertinent information into a standardized location.

## 4 Areas for Further Work

We believe that the proposed standardized DICOM object and accompanying mathematical models will serve as a starting point to facilitate research on dose forgiveness, both intra- and inter-institutionally. However, further research is needed to address limitations in the characterization of organ functional capacity and dose mapping.

While the distinction between parallel and serial organs can be made in the software, the algorithm can be improved so that the effect of re-irradiation can be more accurately measured. Dose accumulation calculations to approximate dose toxicities from previous radiation are more useful for serial organs than parallel organs. Most of the restrictions to re-irradiation are founded in the prior dose serial organs were exposed to. For these serial organs, prior dose maps will be processed with forgiveness algorithms. In the case of parallel organs, the software should instead calculate the prior volume exposed to radiation through dose map analysis. There are no studies that support dose adjustment based on the previously radiated volume, but prior radiated volume will be shown in the software to clinicians to help gauge the potential reserve capacity of the organ. But understanding a patient’s functional reserve can be challenging and often rely on both objective measures (like PFTs) and also subjective measures like symptoms on exertion. This information can likely be found in the EMR, differing in a per-institution basis. Additionally, functional reserve metrics change from organ to organ; measures to gauge functional lung will be different from measures for functional liver or kidney. In the future, we conceive even this functional information will be included in the DICOM so that subsequent radiation courses can take into account patient functional status for parallel organs. There are also several technical challenges that must be solved to enable the new standard. The most important is determining how to reliably map RT doses onto the reference model such that the voxels at each distinct time point correspond to the same underlying tissue, a process known as dose warping. This is currently accomplished through the use of deformable image registration (DIR), a mathematical operation that creates a transformation which maximizes similarity between two images. In general, we choose one image as the reference point and map other images onto it. While we have made excellent progress in DIR, there remain significant limitations to DIR techniques: transformations are most accurate in regions with high contrast and unique landmarks. These features serve as reference points to ensure that the transformation is accurately warping the entirety of the source image. Low contrast or homogeneous regions like the lung and prostate can result in many solutions that yield the same similarity score with no clear ground-truth. Furthermore, uncertainty in dose warping sharply increases in situations where this is not a one to one mapping between voxels in the reference and source image^26,27^. Recent advancements in using artificial intelligence algorithms to perform image registration showed some promising results but a robust method has not yet been developed. Organs routinely appear to shrink and expand between images, likely due to daily variation, weight loss/gain, or tumor response to radiation. Major anatomical changes, such as a radical resection, provide even more of a challenge. Even within one RT session, we cannot assume that anatomy is static. For example, there exist several techniques to compensate for the effects of respiratory motion on lung tumor volume, including free-breathing, tracking, and gating^28^. To mitigate this challenge, we propose keeping track of these deformations in a separate file, this will enable software to re-calculate the un-deformed dose maps delivered during prior radiation so that clinicians can choose the type of deformation they are comfortable with.

Another important challenge to solve is ensuring that there is interoperability between radiation planning software packages, including those from MIM Software, RaySearch Laboratories, and Varian Medical Systems. Clinicians at a hospital that uses MIM Maestro© should be able to open a new patient’s medical record and seamlessly import a reference DICOM file from the previous treatment team generated with RayStation©. Verifying interoperability may come in the form of a test suite, created by a consortium of RT software companies, which is run before the release of every new product version. The details of the test suite are beyond the scope of this proposal but would draw inspiration from similar efforts such as the IHE-RO initiative^29^. Alternatively, information about data formatting could be made part of the standard, thereby ensuring that data adheres to a format in a manufacturer-agnostic way.

Finally, we anticipate that each patient will have unique tissue recovery parameters. In order to further personalize radiation courses, work should be conducted on studying how patient genomics can be used to adjust the models detailed in prior sections, much like how genomic-adjusted radiation dose (GARD) has been shown to predict tumor response to radiotherapy^30,31^, and allow for formal optimization combining normal tissue and tumor control models.^32^

While these challenges represent important limitations in our proposed standard, we believe there are compelling immediate potential improvements in both clinical care and long term re-irradiation dose modeling from adoption of our proposed model.

## 5 Conclusion

Integrating treatment information into a single DICOM file offers substantial advantages that enhance both clinical practice and research. Initially, this approach will facilitate research on dose forgiveness by leveraging patient outcomes from repeat radiation courses. It will also allow radiation oncologists to evaluate various dose forgiveness algorithms from prior radiation plans, on an organ-by-organ basis, and make more informed decisions about future radiation. In the future, even basic side-effect profiles can be included so clinicians can better understand dose tolerance and tissue recovery between treatments. This analysis, while already possible with direct EMR access, becomes more versatile when incorporated into DICOM, enabling use across various software applications like treatment planning and contouring aids.

Embedding this data within DICOM files promotes inter-institutional research and analysis. When patients receive treatments at different institutions, technological constraints often limit access to comprehensive treatment details. Including this information in DICOM files eliminates the need for separate record transfers, ensuring continuity and completeness of data. Moreover, recording treatment side-effects within DICOM files supports intra-institutional tracking of treatment tolerance and dose forgiveness, thereby fostering collaborative research efforts and enabling the pooling of patient data for more precise analyses.

## Data Availability

Data will be provided upon request.

## References

1. Katipally, R. R., Pitroda, S. P., Juloori, A., Chmura, S. J. & Weichselbaum, R. R. The oligometastatic spectrum in the era of improved detection and modern systemic therapy. Nat. Rev. Clin. Oncol. 19, 585–599, DOI: 10.1038/s41571-022-00655-9 (2022).

2. Palma, D. A. et al. Stereotactic ablative radiotherapy versus standard of care palliative treatment in patients with oligometastatic cancers (SABR-COMET): a randomised, phase 2, open-label trial. The Lancet 393, 2051–2058, DOI: 10.1016/s0140-6736(18)32487-5 (2019).

3. Gomez, D. R. et al. Local consolidative therapy vs. maintenance therapy or observation for patients with oligometastatic non–small-cell lung cancer: Long-term results of a multi-institutional, phase II, randomized study. J. Clin. Oncol. 37, 1558–1565, DOI: 10.1200/jco.19.00201 (2019).

4. Timmerman, R. A story of hypofractionation and the table on the wall. Int. J. Radiat. Oncol. Biol. Phys. 112, 4–21 (2022).

5. Daly, M. E. et al. Normal tissue complication probability estimation by the Lyman-Kutcher-Burman method does not accurately predict spinal cord tolerance to stereotactic radiosurgery. Int. J. Radiat. Oncol. * Biol. * Phys. 82, 2025–2032, DOI: 10.1016/j.ijrobp.2011.03.004 (2012).

6. Dennstädt, F., Medová, M., Putora, P. M. & Glatzer, M. Parameters of the lyman model for calculation of normal-tissue complication probability: A systematic literature review. Int. J. Radiat. Oncol. Biol. Phys. 115, 696–706, DOI: 10.1016/j.ijrobp.2022.08.039 (2023).

7. Jones, B. & Hopewell, J. W. Alternative models for estimating the radiotherapy retreatment dose for the spinal cord. Int. J. Radiat. Biol. 90, 731–741, DOI: 10.3109/09553002.2014.925151 (2014).

8. Paradis, K. et al. The special medical physics consult process for reirradiation patients. Adv Radiat Oncol 4, 559–565, DOI: 10.1016/j.adro.2019.05.007 (2019).

9. Sahgal, A. et al. Spinal cord dose tolerance to stereotactic body radiation therapy. Int. J. Radiat. Oncol. * Biol. * Phys. DOI: 10.1016/j.ijrobp.2019.09.038 (2019).

10. Scarborough, J. A., Tom, M. C., Kattan, M. W. & Scott, J. G. Revisiting a null hypothesis: Exploring the parameters of oligometastasis treatment. Int. journal radiation oncology, biology, physics 110, 371–381, DOI: 10.1016/j.ijrobp.2020.12.044 (2021).

11. Beckham, T. H., Yang, T. J., Gomez, D. & Tsai, C. J. Metastasis-directed therapy for oligometastasis and beyond. Br. J. Cancer 124, 136–141, DOI: 10.1038/s41416-020-01128-5 (2021).

12. Kerns, S. L., Ostrer, H. & Rosenstein, B. S. Radiogenomics: using genetics to identify cancer patients at risk for development of adverse effects following radiotherapy. Cancer discovery 4, 155–165 (2014).

13. Lee, J., Kim, C. Y., Koom, W. S. & Rim, C. H. Practical effectiveness of re-irradiation with or without surgery for locoregional recurrence of rectal cancer: A meta-analysis and systematic review. Radiother. Oncol. 140, 10–19, DOI: 10.1016/j.radonc.2019.05.021 (2019).

14. Smith, T. et al. Stereotactic body radiation therapy reirradiation for locally recurrent rectal cancer: Outcomes and toxicity. Adv. Radiat. Oncol. 5, 1311–1319, DOI: 10.1016/j.adro.2020.07.017 (2020).

15. Dörr, W. & Gabryş, D. The principles and practice of re-irradiation in clinical oncology: An overview. Clin. Oncol. 30, 67–72, DOI: 10.1016/j.clon.2017.11.014 (2018).

16. Parker, C. C. et al. Radiotherapy to the primary tumour for newly diagnosed, metastatic prostate cancer (STAMPEDE): a randomised controlled phase 3 trial. The Lancet 392, 2353–2366 (2018).

17. Ruers, T. et al. Local treatment of unresectable colorectal liver metastases: results of a randomized phase II trial. JNCI: J. Natl. Cancer Inst. 109, djx015 (2017).

18. Ost, P. et al. Surveillance or metastasis-directed therapy for oligometastatic prostate cancer recurrence: a prospective, randomized, multicenter phase II trial. J. Clin. Oncol. 36, 446–453 (2018).

19. Ang, K. et al. The tolerance of primate spinal cord to re-irradiation. Int. J. Radiat. Oncol. * Biol. * Phys. 25, 459–464, DOI: 10.1016/0360-3016(93)90067-6 (1993).

20. Myrehaug, S., Soliman, H., Tseng, C., Heyn, C. & Sahgal, A. Re-irradiation of vertebral body metastases: Treatment in the radiosurgery era. Clin. Oncol. 30, 85–92, DOI: 10.1016/j.clon.2017.11.005 (2018).

21. Hashmi, A. et al. Re-irradiation stereotactic body radiotherapy for spinal metastases: a multi-institutional outcome analysis. J. Neurosurgery: Spine 25, 646 – 653, DOI: 10.3171/2016.4.SPINE151523 (2016).

22. Association, N. E. M. Digital imaging and communications in medicine (dicom) standard (2023).

23. DICOM Standards Committee, Working Group 07. Supplement 147: Second Generation Radiotherapy (2018).

24. Xie, Y.-H.Chen, Y.-X. & Fang, J.-Y. Comprehensive review of targeted therapy for colorectal cancer. Signal Transduct. Target. Ther. 5, 1–30, DOI: 10.1038/s41392-020-0116-z (2020). Publisher: Nature Publishing Group.

25. Herbst, R. S. et al. COAST: An open-label, phase II, multidrug platform study of durvalumab alone or in combination with oleclumab or monalizumab in patients with unresectable, stage III non-small-cell lung cancer. J. Clin. Oncol. Off. J. Am. Soc. Clin. Oncol. 40, 3383–3393, DOI: 10.1200/JCO.22.00227 (2022).

26. Motegi, K. et al. Usefulness of hybrid deformable image registration algorithms in prostate radiation therapy. J. Appl. Clin. Med. Phys. 20, 229–236, DOI: 10.1002/acm2.12515 (2019).

27. Qin, A., Liang, J., Han, X., O’Connell, N. & Yan, D. Technical note: The impact of deformable image registration methods on dose warping. Med. Phys. 45, 1287–1294, DOI: 10.1002/mp.12741 (2018).

28. Prunaretty, J. et al. Tracking, gating, free-breathing, which technique to use for lung stereotactic treatments? a dosimetric comparison. Reports Pract. Oncol. Radiother. 24, 97–104 (2019).

29. Rengan, R. et al. Addressing connectivity issues: The integrating the healthcare enterprise-radiation oncology (IHE-RO) initiative. Pract. radiation oncology 1, 226–231 (2011).

30. Scott, J. G. et al. A genome-based model for adjusting radiotherapy dose (GARD): a retrospective, cohort-based study. The lancet oncology 18, 202–211 (2017).

31. Scott, J. G. et al. Pan-cancer prediction of radiotherapy benefit using genomic-adjusted radiation dose (GARD): a cohort-based pooled analysis. The Lancet Oncol. 22, 1221–1229 (2021).

32. Scott, J. G. et al. Personalizing radiotherapy prescription dose using genomic markers of radiosensitivity and normal tissue toxicity in nsclc. J. Thorac. Oncol. 16, 428–438 (2021).

